# Classification of Omicron BA.1, BA.1.1 and BA.2 sublineages by TaqMan assay consistent with whole genome analysis data

**DOI:** 10.1101/2022.04.03.22273268

**Authors:** Yosuke Hirotsu, Makoto Maejima, Masahiro Shibusawa, Yume Natori, Yuki Nagakubo, Kazuhiro Hosaka, Hitomi Sueki, Hitoshi Mochizuki, Toshiharu Tsutsui, Yumiko Kakizaki, Yoshihiro Miyashita, Masao Omata

## Abstract

**Objective:** Recently, the Omicron strain of severe acute respiratory syndrome coronavirus 2 (SARS-CoV-2) has spread and replaced the previously dominant Delta strain. Several Omicron sublineages (BA.1, BA.1.1 and BA.2) have been identified, with in vitro and preclinical reports showing that the pathogenicity and therapeutic efficacy differs between BA.1 and BA.2. We sought to develop a TaqMan assay to identify these subvariants.

**Methods:** A TaqMan assay was constructed for rapid identification and genotyping of Omicron sublineages. We analyzed three characteristic mutations of the spike gene, Δ69–70, G339D and Q493R, by TaqMan assay. The accuracy of the TaqMan assay was examined by comparing its results with the results of whole genome sequencing (WGS) analysis.

**Results:** A total of 169 SARS-CoV-2 positive samples were analyzed by WGS and TaqMan assay. The 127 samples determined as BA.1/BA.1.1 by WGS were all positive for Δ69–70, G339D and Q493R by TaqMan assay. Forty-two samples determined as BA.2 by WGS were negative for Δ69–70 but positive for G339D and Q493R by TaqMan. The concordance rate between WGS and the TaqMan assay was 100% (169/169).

**Conclusion:** TaqMan assays targeting characteristic mutations are useful for identification and discrimination of Omicron sublineages.

## Introduction

The global pandemic of severe acute respiratory syndrome coronavirus 2 (SARS-CoV-2) has become a threat to human life. However, since its outbreak, scientific knowledge has accumulated on the transmission and pathogenicity of this virus. Additionally, the development of vaccines and antiviral treatments has contributed to preventing infection and reduced the risk of severe illness and mortality.

SARS-CoV-2 is an RNA virus with a genome size of approximately 30 kb [1]. SARS-CoV-2 mutates at a rate of approximately two mutations per month [2], and is prone to mutations most frequently occurring in the *spike* and *orf1ab* genes [3]. These mutations can affect transmissibility of the virus and efficacy of antiviral drugs and vaccines. To control the emergence of new strains, it is important to continuously monitor viral evolution through genomic epidemiological analysis [4].

Recently, a new strain designated B.1.1.529 was detected and isolated in Gauteng Province, South Africa [5]. The World Health Organization designated the B.1.1.529 lineage as a variant of concern (VOC) on November 26, 2021 [5], naming it Omicron in line with the naming convention adopted for previous VOCs using the Greek alphabet, and it has since spread to a number of countries to become a globally dominant strain [6]. Omicron contains more than 30 nonsynonymous mutations in the spike protein, therefore Omicron viruses may reduce the efficacy of antibody therapy and evade vaccine-induced immunity.

Omicron viruses are classified into several sublineages including BA.1, BA.1.1, BA.2 and BA.3, which have been observed worldwide. In the early period following Omicron emergence, BA.1 was the dominant sublineage; however in Denmark, the United Kingdom, India, the Philippines, and South Africa, BA.2 became dominant [7-9], with BA.2 reported to be more transmissible than BA.1 [10]. Convalescent sera and sera from mRNA vaccine recipients showed decreased neutralizing activity against BA.1, BA,1.1. and BA.2 [11-13]. In infection experiments using hamsters, BA.2 caused more inflammation in the lungs and was more pathogenic than BA.1 [8]. Antibody cocktail therapy (casirivimab-imdevimab), which had lost neutralizing activity in BA.1 and BA.1.1, showed partially neutralizing activity in BA.2 [14]. In addition, S309 (sotrovimab) demonstrated less neutralization activity against BA.2 compared with BA.1 and BA.1.1 [14, 15]. The RNA-dependent RNA polymerase inhibitors, remdesivir and molnupiravir, and the protease inhibitor nirmatrelvir remained effective in BA.2 as well as BA.1 [14]. Therefore, genotyping of Omicron sublineages is important for determining infection control and treatment strategies.

In this study, we present a TaqMan assay to discriminate between BA.1/BA.1.1 and BA.2. This method is feasible for any laboratory equipped to perform quantitative real-time PCR.

## Methods

### SARS-CoV-2 diagnostic testing

Nasopharyngeal swab samples were collected using cotton swabs and placed in viral transport media (Copan Diagnostics, Murrieta, CA). Multiple molecular diagnostic testing platforms, including SARS-CoV-2 quantitative reverse transcriptase PCR in accordance with the protocol developed by the National Institute of Infectious Diseases in Japan [16], FilmArray Respiratory Panel 2.1 with the FilmArray Torch system (bioMérieux, Marcy-l ‘Etoile, France) [17], Xpert Xpress SARS-CoV-2 test using Cepheid GeneXpert (Cepheid, Sunnyvale, CA) [18] and Lumipulse antigen test with the LUMIPULSE G600II system (Fujirebio, Inc., Tokyo, Japan) were used for this study [19, 20].

### Whole genome sequencing (WGS)

We subjected 171 samples collected from coronavirus disease 2019 (COVID-19) patients to whole genome analysis (Table S1). WGS analysis was performed as previously described [21-23]. In brief, SARS-CoV-2 genomic RNA was reverse transcribed into cDNA and amplified by using the Ion AmpliSeq SARS-CoV-2 Insight Research Assay (Thermo Fisher Scientific, Waltham MA) on the Ion Torrent Genexus System. Sequencing reads and quality were processed using Genexus software with SARS-CoV-2 plugins. The sequencing reads were mapped and aligned using the torrent mapping alignment program. Assembly was performed with the Iterative Refinement Meta-Assembler [24].

To determine the viral clade and lineage classifications, the consensus FASTA files were downloaded and processed through Nextstrain [25], and Phylogenetic Assignment of Named Global Outbreak Lineages (PANGOLIN) [26]. The FASTA files were deposited in the Global Initiative on Sharing Avian Influenza Data (GISAID) EpiCoV database [2]. All GISAID Accession IDs are noted in Table S1.

### TaqMan assay

We used the pre-designed TaqMan SARS-CoV-2 Mutation Panel for detecting SARS-CoV-2 spike Δ69–70, G339D and Q493R (Thermo Fisher Scientific). The TaqMan probe detected both wild-type and variant sequences of SARS-CoV-2, with the TaqMan minor groove binder probe for the wild-type allele labelled with VIC dye and the variant allele with FAM dye fluorescence. TaqPath 1-Step RT-qPCR Master Mix CG was used as master mix and the qPCR was performed on the Step-One Plus Real Time PCR System (Thermo Fisher Scientific).

### Ethics approval

The Institutional Review Board of the Clinical Research and Genome Research Committee at Yamanashi Central Hospital approved this study and the use of an opt-out consent method (Approval No. C2019-30). The requirement for written informed consent was waived owing to the observational nature of this study and the urgent need to collect COVID-19 data.

## Results

### Multiple mutations are present in Omicron

Omicron contains numerous mutations compared with other VOCs [27-29]. Thirty-three mutations have been identified in the spike protein in BA.1, 34 in BA.1.1 and 29 in BA.2. BA.1 and BA.1.1 share 33 mutations, but BA.1.1 has an additional mutation in R346K compared with BA.1 (Figure 1). Compared with BA.1 and BA.1.1, the spike protein mutations in BA.2 are very different. Unique mutations in BA.1/BA.1.1 are A67V, Δ69–70, T95I, Δ143– 145, N211I, Δ212, S371L, G466S, G496S, T547K, N856K and L981F. Conversely, unique mutations in BA.2 are T19I, L24S, Δ25–27, V213G, S371F, T376A, D405N and R408S.

**Figure 1.**
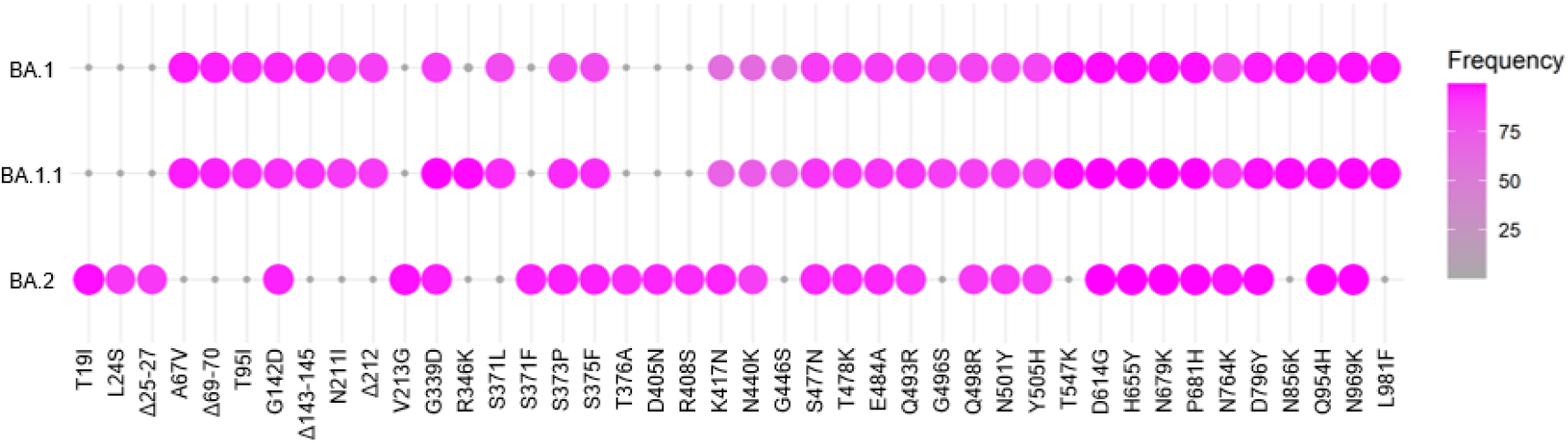
Mutations in Omicron sublineages. Mutations are shown in Omicron sublineages BA.1, BA.1.1 and BA.2. The color gradient indicates the frequency of detected mutations. The pink circles indicate the presence of mutations. The frequency of mutation data was referenced from the website outbreak.info [29].

### Construction of a TaqMan assay to distinguish Omicron sublineages

To distinguish BA.1/BA.1.1 from BA.2, we constructed a TaqMan assay system to analyze the spike protein mutations that characterize the Omicron sublineages. Thus, we used pre-designed TaqMan probes to distinguish Δ69–70, G339D and Q493R. Spike G339D and Q493R mutations are detected in BA.1/BA.1.1 and BA.2 but not in other VOCs (Figure S1). The spike Δ69–70 mutation was also used to distinguish sublineages because it is detected in BA.1/BA.1.1 but not in BA.2 (Figure 1). When analyzed using nucleic acids extracted from nasopharyngeal swabs, G339D and Q493R mutations were specifically detected in Omicron-infected patients (Figure 2A and 2B). In addition, the Δ69–70 mutation was distinct between BA1/BA.1.1 and BA.2 (Figure 2C). These results showed that TaqMan assays could detect Omicron strains and distinguish between Omicron sublineages.

**Figure 2.**
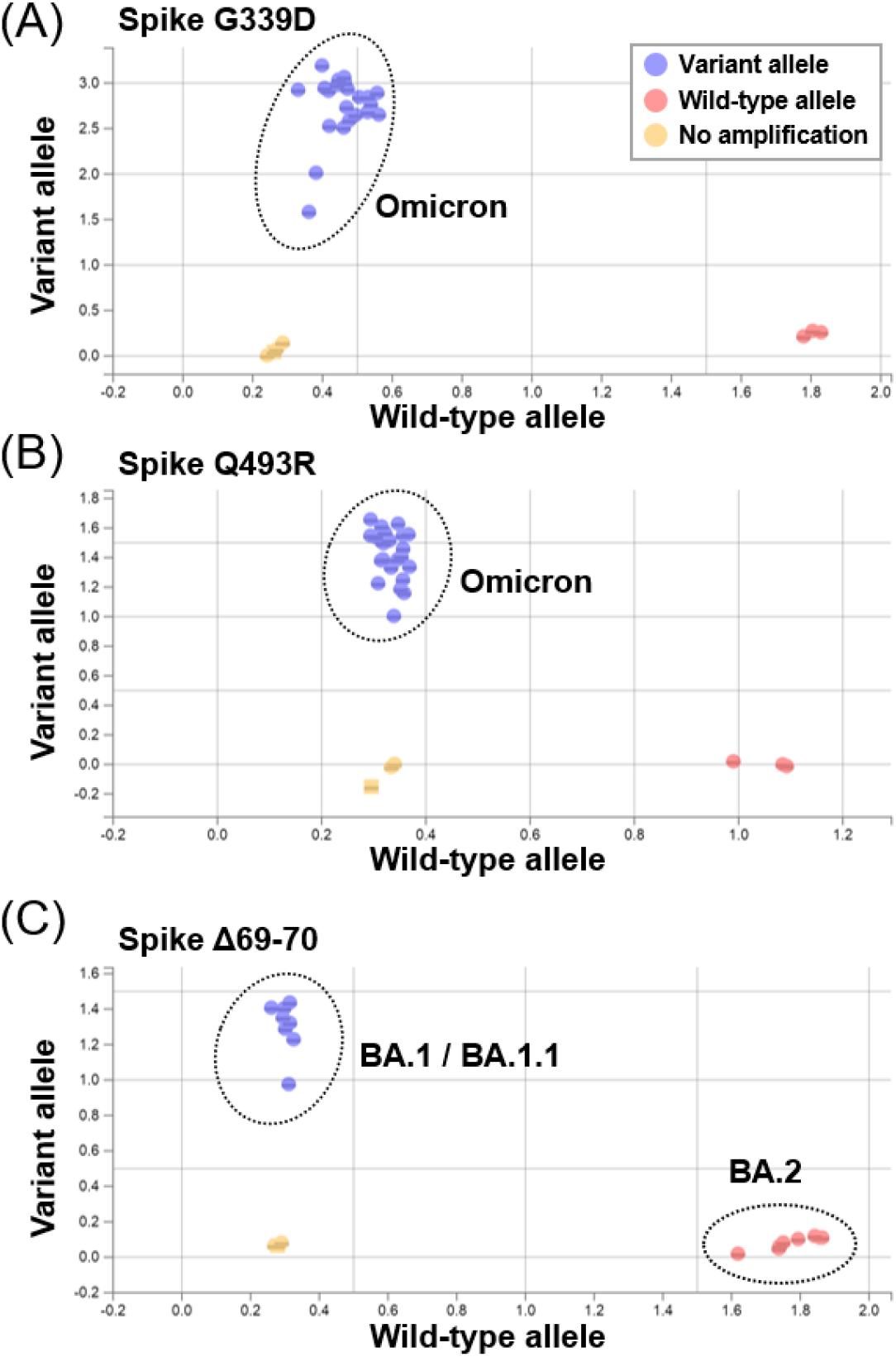
Genotyping of Omicron sublineages by TaqMan assay. **(A-C)** Samples were analyzed with a TaqMan assay that detects mutations in Omicron spike proteins. Spike protein mutations G339D (A), Q493R (B) and Δ69–70 (C) were targeted. G399D and Q493R indicated Omicron (including BA.1/BA.1.1 and BA.2), while Δ69–70 was used to distinguish BA.1/BA.1.1 from BA.2. Blue circles indicate variant alleles (FAM dye) and red circles indicate wild-type alleles (VIC dye).

### Comparison of TaqMan assay and WGS data

To examine whether the TaqMan assay could accurately distinguish Omicron sublineages, we compared WGS data and TaqMan assay results using 169 SARS-CoV-2 positive samples (Table 1). WGS analysis determined 127 samples to be BA.1/BA.1.1 and 42 samples to be BA.2 (Table 1). In these samples, TaqMan assay analysis showed that all BA.1/BA.1.1 samples were positive for Δ69–70, G339D and Q493R, while BA.2 samples were negative for Δ69–70 and positive for G339D and Q493R (Table 1). The TaqMan assay data were consistent with WGS data, demonstrating 100% (169/169) agreement. Therefore, the TaqMan assay represents a useful technique for distinguishing Omicron sublineages.

**Table 1.**
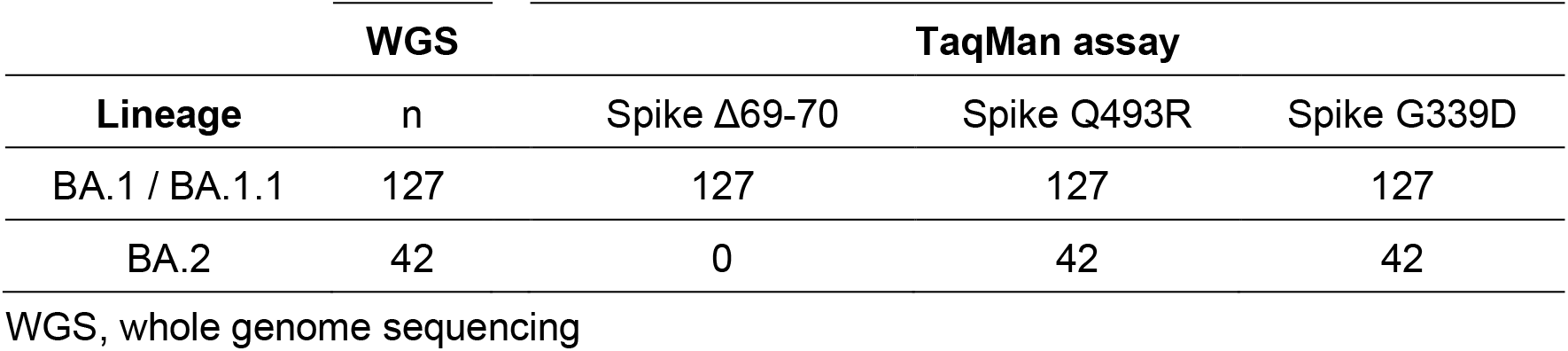
Comparison of results between WGS and TaqMan assay.

### Mutations in TaqMan assay target sites

During WGS analysis of serially collected samples, SARS-CoV-2 viruses with the spike G339N mutation (position 22577–22578: c.1015_1016delGGGinsAA) were detected in two individuals (Accession ID: EPI_ISL_11018144 and EPI_ISL_11018145) (Figure 3A). Analysis with PANGOLIN classified these samples as BA.2. The mutation that occurred at codon 399 overlapped with the target site of the TaqMan assay. We therefore examined whether the TaqMan assay targeting G339D could efficiently amplify the target site in the sample with G399N. In both BA.2 G339D and BA.2 G339N samples, sufficient amplification signals were obtained for Δ69–70 and Q493R. However, compared with BA.2 G339D, BA.2 G339N showed a lower amplification efficiency of the variant allele-specific signal (blue line in Figure 3B). In conclusion, the TaqMan assay used in this study specifically detected characteristic mutations related to Omicron sublineages in clinical samples.

**Figure 3.**
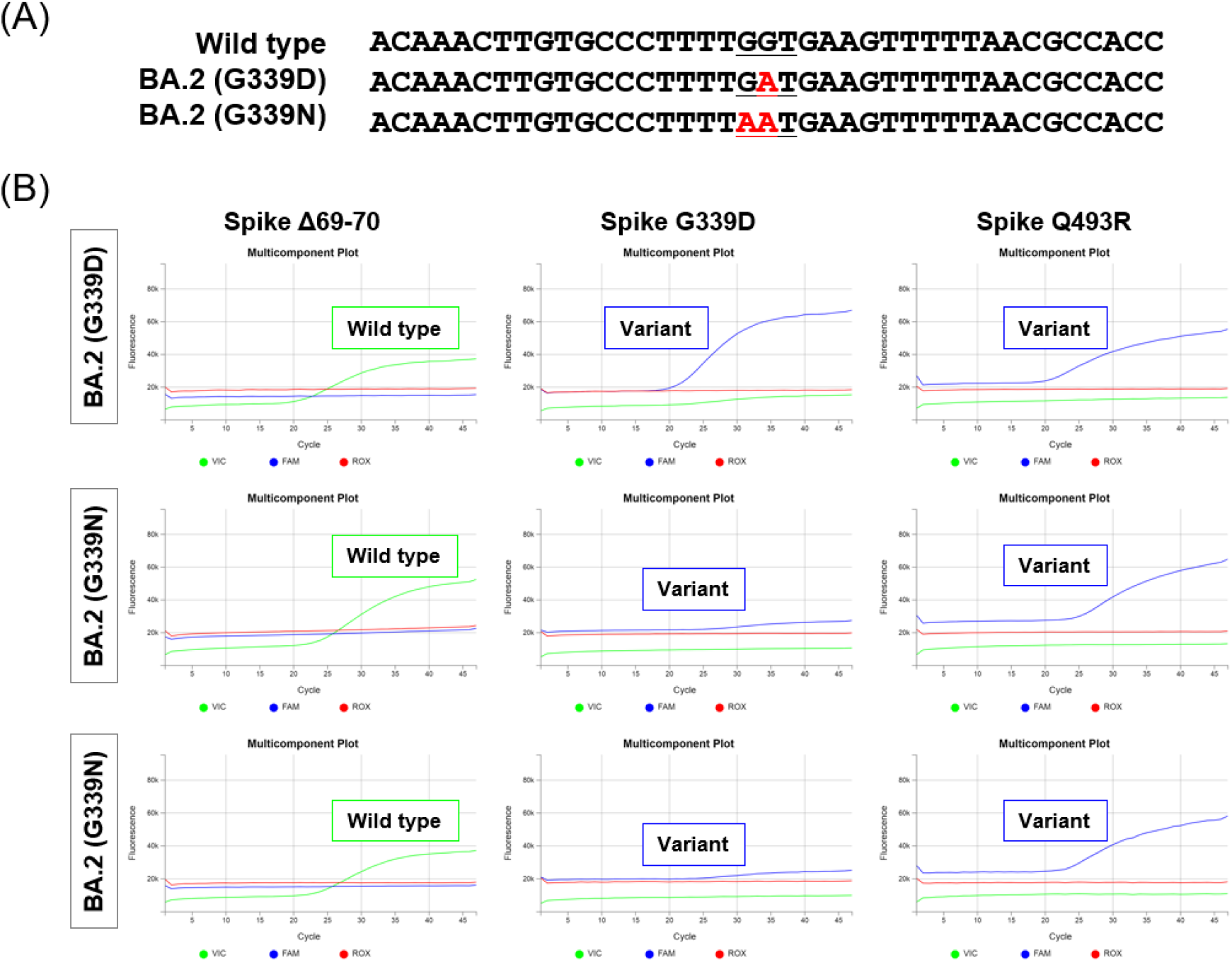
Analysis of samples with mutations in the TaqMan probe site. Two samples were classified by WGS analysis as BA.2 containing a G339N mutation. **(A)** The mutation site occurred at codon 339, from wild type (GGT, glycine [G]) to the variant form (GAT, aspartate [D] or AAT, aspartate [N]). **(B)** Most BA.2 lineage viruses carry the G339D mutation, but two samples showed a G339N mutation by WGS. Compared with BA.2 G339D, the fluorescent signal (blue line) was lower in the BA.2 G339N samples.

## Discussion

In this study, we present data regarding the use of a TaqMan assay to distinguish between Omicron strains and their sublineages. WGS analysis is the most standard method to determine these sublineages. However, WGS analysis of all specimens is restricted by limited resources and is difficult to apply under conditions of rapid spread of infection. In this regard, we have established a TaqMan assay that more conveniently and rapidly identifies Omicron strains and distinguishes the sublineages. BA.1/BA.1.1 and BA.2 are reported to differ in transmissibility and treatment response, and so the World Health Organization recommends monitoring BA.2 as a separate sublineage [30]. Therefore, assay systems that distinguish these subtypes will be important in determining preventive measures, infection control and treatment strategies.

Each viral lineage has its own characteristic mutations. By targeting these mutations, it is possible to distinguish mutant strains and sublineages. Based on the accumulating WGS data during surveillance, we consider the TaqMan assay to be a suitable approach to detect characteristic mutations that occur frequently among viral lineages. According to the GISAID database, as of March 11, 2022, the frequency of mutations targeted in this study detected in each sublineage were as follows. Δ69-70 in 96.1%, 95.7% and 0.1% of BA.1, BA.1.1 and BA.2, respectively; G339D in 87.9%, 99.3% and 96.3% of BA.1, BA.1.1 and BA.2, respectively; Q493R in 88.3%, 91.2% and 92% of BA.1, BA.1.1 and BA.2, respectively [29]. It is also possible to target other mutations as reported by other groups [31, 32]. Analysis of multiple characteristic mutations is expected to improve the accuracy of classification of virus lineage.

WGS analysis generally requires labor-intensive procedures such as reverse transcription reactions from RNA to cDNA, library preparation, purification and library quantification. In addition, facilities and laboratories with limited analytical equipment and resources cannot perform WGS analysis. Alternatively, TaqMan assays are useful to distinguish VOCs and their sublineages quickly and easily in any laboratory with qPCR facilities. Recently, a Delta-Omicron hybrid strain (AY.4/BA.1 recombinant, EPI_ISL_10819657) was reported in France and has been detected in several European countries [2, 33]. The TaqMan method could be adapted to identify any such newly emerging mutant strains.

There are, however, several limitations. TaqMan assays do not always accurately determine the sublineages in some situations. WGS analysis may be necessary when the characteristic mutations of newly-emerging strains or sublineages remain under investigation. It should also be noted that if mutations occur at the TaqMan probe or primer locations, the PCR amplification efficiency will be reduced and the signal will be attenuated, which may make interpretation difficult (Figure 3B).

Our study showed that the TaqMan assay can be used to specifically detect Omicron viruses and classify subvariants. By applying this method to SARS-CoV-2-positive specimens, it is possible to analyze multiple specimens rapidly. Although accumulated data are still needed [8, 11, 14, 34], BA.1 and BA.2 show different susceptibilities to antibody and antiviral therapy. Rapid classification of Omicron sublineages may be clinically critical in providing appropriate treatment to patients with COVID-19.

## Data Availability

The sequences of SARS-CoV-2 genomes are available on GISAID (www.gisaid.org). We have provided the accession numbers in the Supplementary Information. Source data are provided with this paper.

## Acknowledgements

We thank all medical and ancillary hospital staff for their support. We thank Gillian Campbell, PhD, from Edanz (https://www.jp.edanz.com/ac) for editing a draft of this manuscript.

---

**Figure S1.**
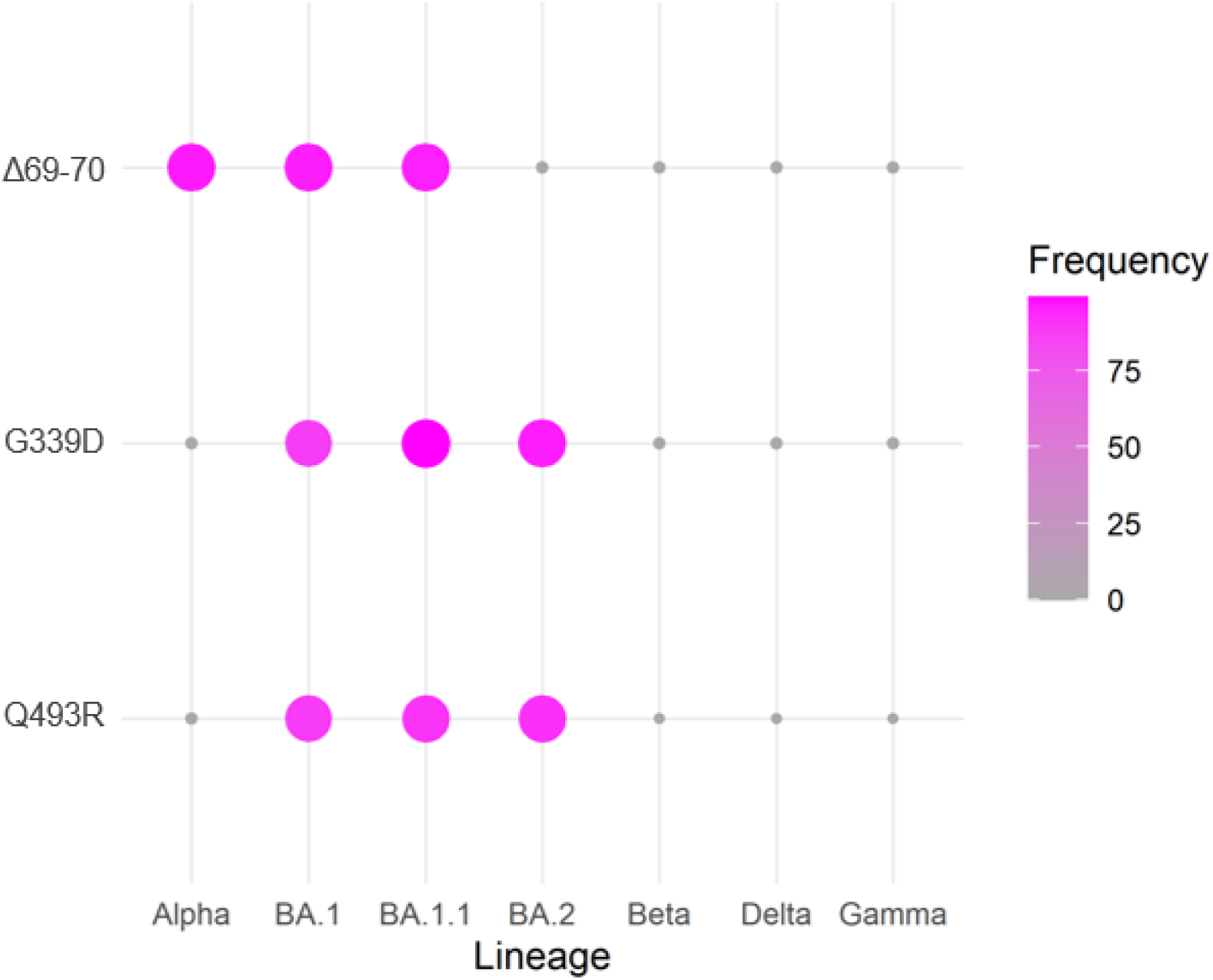
Mutation frequency in VOCs and Omicron sublineages. The percentages of spike mutations Δ69–70, G339D and Q493R in each VOC analyzed in this study are shown. The pink circles indicate the presence of mutations. The frequency of mutation data was referenced from the website outbreak.info [29].

