## Supplemental Table 1 for "Classification of Omicron BA.1, BA.1.1 and BA.2 sublineages by TaqMan assay consistent with whole genome analysis data"

**Supplemental Table 1. List of results for whole genome sequencing analysis and TaqMan assay**

| **Case #** | **Age** | **Sex** | **GISAID Accession ID** | **Lineage determined by WGS** | **Spike ΔH69V70** | **Spike Q493R** | **Spike G339D** |
| --- | --- | --- | --- | --- | --- | --- | --- |
| 1 | 6-10 | Male | EPI_ISL_9203995 | BA.1.1 | Positive | Positive | Positive |
| 2 | 16-20 | Female | EPI_ISL_9203996 | BA.1.1 | Positive | Positive | Positive |
| 3 | 21-25 | Male | EPI_ISL_9203997 | BA.1.1 | Positive | Positive | Positive |
| 4 | 56-60 | Male | EPI_ISL_9203998 | BA.1.1 | Positive | Positive | Positive |
| 5 | 31-35 | Female | EPI_ISL_9203999 | BA.1.1 | Positive | Positive | Positive |
| 6 | 16-20 | Male | EPI_ISL_9204000 | BA.1 | Positive | Positive | Positive |
| 7 | 16-20 | Male | EPI_ISL_9204001 | BA.1.1 | Positive | Positive | Positive |
| 8 | 36-40 | Female | EPI_ISL_9204002 | BA.1.1 | Positive | Positive | Positive |
| 9 | 0-5 | Male | EPI_ISL_9204003 | BA.1.1 | Positive | Positive | Positive |
| 10 | 0-5 | Female | EPI_ISL_9204004 | BA.1.1 | Positive | Positive | Positive |
| 11 | 0-5 | Male | EPI_ISL_9204005 | BA.1.1 | Positive | Positive | Positive |
| 12 | 0-5 | Male | EPI_ISL_9204006 | BA.1.1 | Positive | Positive | Positive |
| 13 | 16-20 | Female | EPI_ISL_9204007 | BA.1.1 | Positive | Positive | Positive |
| 14 | 26-30 | Male | EPI_ISL_9204009 | BA.1.1 | Positive | Positive | Positive |
| 15 | 16-20 | Male | EPI_ISL_9375180 | BA.1.1 | Positive | Positive | Positive |
| 16 | 36-40 | Male | EPI_ISL_9375181 | BA.1.1 | Positive | Positive | Positive |
| 17 | 26-30 | Female | EPI_ISL_9375182 | BA.1.1 | Positive | Positive | Positive |
| 18 | 16-20 | Female | EPI_ISL_9375183 | BA.1.1 | Positive | Positive | Positive |
| 19 | 26-30 | Female | EPI_ISL_9375184 | BA.1.1 | Positive | Positive | Positive |
| 20 | 16-20 | Female | EPI_ISL_9375185 | BA.1.1 | Positive | Positive | Positive |
| 21 | 16-20 | Male | EPI_ISL_9375186 | BA.1.1 | Positive | Positive | Positive |
| 22 | 16-20 | Female | EPI_ISL_9375187 | BA.1.1 | Positive | Positive | Positive |
| 23 | 56-60 | Female | EPI_ISL_9413709 | BA.1.1 | Positive | Positive | Positive |
| 24 | 56-60 | Female | EPI_ISL_9413710 | BA.1.1 | Positive | Positive | Positive |
| 25 | 36-40 | Male | EPI_ISL_9413711 | BA.1.1 | Positive | Positive | Positive |
| 26 | 16-20 | Male | EPI_ISL_9413712 | BA.1.1 | Positive | Positive | Positive |
| 27 | 96-100 | Female | EPI_ISL_9413713 | BA.1.1 | Positive | Positive | Positive |
| 28 | 46-50 | Male | EPI_ISL_9413714 | BA.1.1 | Positive | Positive | Positive |
| 29 | 16-20 | Male | EPI_ISL_9579097 | BA.1.1 | Positive | Positive | Positive |
| 30 | 46-50 | Male | EPI_ISL_9579098 | BA.1.1 | Positive | Positive | Positive |
| 31 | 41-45 | Female | EPI_ISL_9579099 | BA.1.1 | Positive | Positive | Positive |
| 32 | 81-85 | Female | EPI_ISL_9579100 | BA.1.1 | Positive | Positive | Positive |
| 33 | 21-25 | Male | EPI_ISL_9579101 | BA.1.1 | Positive | Positive | Positive |
| 34 | 51-55 | Male | EPI_ISL_9579102 | BA.1.1 | Positive | Positive | Positive |
| 35 | 16-20 | Male | EPI_ISL_9579110 | BA.1.1 | Positive | Positive | Positive |
| 36 | 51-55 | Male | EPI_ISL_9596561 | BA.1.1 | Positive | Positive | Positive |
| 37 | 11-15 | Male | EPI_ISL_9596562 | BA.1.1 | Positive | Positive | Positive |
| 38 | 61-65 | Female | EPI_ISL_9596563 | BA.1.1 | Positive | Positive | Positive |
| 39 | 91-95 | Male | EPI_ISL_9596564 | BA.1.1 | Positive | Positive | Positive |
| 40 | 76-80 | Male | EPI_ISL_9596565 | BA.1.1 | Positive | Positive | Positive |
| 41 | 96-100 | Female | EPI_ISL_9596566 | BA.1.1 | Positive | Positive | Positive |
| 42 | 96-100 | Female | EPI_ISL_9596568 | BA.1.1 | Positive | Positive | Positive |
| 43 | 31-35 | Male | EPI_ISL_9596569 | BA.1.1 | Positive | Positive | Positive |
| 44 | 6-10 | Male | EPI_ISL_9596570 | BA.1.1 | Positive | Positive | Positive |
| 45 | 66-70 | Female | EPI_ISL_9596571 | BA.1.1 | Positive | Positive | Positive |
| 46 | 21-25 | Female | EPI_ISL_9596572 | BA.1.1 | Positive | Positive | Positive |
| 47 | 26-30 | Female | EPI_ISL_9596573 | BA.1.1 | Positive | Positive | Positive |
| 48 | 41-45 | Male | EPI_ISL_9636551 | BA.1.1 | Positive | Positive | Positive |
| 49 | 91-95 | Male | EPI_ISL_9596574 | BA.1.1 | Positive | Positive | Positive |
| 50 | 41-45 | Male | EPI_ISL_9596575 | BA.1.1 | Positive | Positive | Positive |
| 51 | 26-30 | Male | EPI_ISL_9596576 | BA.1.1 | Positive | Positive | Positive |
| 52 | 66-70 | Female | EPI_ISL_9596577 | BA.1.1 | Positive | Positive | Positive |
| 53 | 21-25 | Female | EPI_ISL_9596578 | BA.1.1 | Positive | Positive | Positive |
| 54 | 16-20 | Male | EPI_ISL_9596580 | BA.1.1 | Positive | Positive | Positive |
| 55 | 16-20 | Male | EPI_ISL_9596581 | BA.1.1 | Positive | Positive | Positive |
| 56 | 21-25 | Male | EPI_ISL_9596582 | BA.1.1 | Positive | Positive | Positive |
| 57 | 21-25 | Female | EPI_ISL_9596583 | BA.1.1 | Positive | Positive | Positive |
| 58 | 91-95 | Female | EPI_ISL_9596584 | BA.1.1 | Positive | Positive | Positive |
| 59 | 0-5 | Male | EPI_ISL_9596585 | BA.1.1 | Positive | Positive | Positive |
| 60 | 76-80 | Male | EPI_ISL_9596586 | BA.1.1 | Positive | Positive | Positive |
| 61 | 46-50 | Male | EPI_ISL_9596587 | BA.1.1 | Positive | Positive | Positive |
| 62 | 36-40 | Female | EPI_ISL_9596588 | BA.1.1 | Positive | Positive | Positive |
| 63 | 16-20 | Female | EPI_ISL_9596589 | BA.1.1 | Positive | Positive | Positive |
| 64 | 0-5 | Male | EPI_ISL_9596590 | BA.1.1 | Positive | Positive | Positive |
| 65 | 26-30 | Male | EPI_ISL_9640042 | BA.1.1 | Positive | Positive | Positive |
| 66 | 16-20 | Female | EPI_ISL_9640043 | BA.1.1 | Positive | Positive | Positive |
| 67 | 46-50 | Female | EPI_ISL_9640044 | BA.1.1 | Positive | Positive | Positive |
| 68 | 66-70 | Male | EPI_ISL_9640045 | BA.1.1 | Positive | Positive | Positive |
| 69 | 66-70 | Female | EPI_ISL_9640046 | BA.1.1 | Positive | Positive | Positive |
| 70 | 51-55 | Male | EPI_ISL_9640047 | BA.1.1 | Positive | Positive | Positive |
| 71 | 66-70 | Female | EPI_ISL_9640048 | BA.1.1 | Positive | Positive | Positive |
| 72 | 21-25 | Male | EPI_ISL_9640049 | BA.1.1 | Positive | Positive | Positive |
| 73 | 26-30 | Female | EPI_ISL_9640050 | BA.1.1 | Positive | Positive | Positive |
| 74 | 61-65 | Male | EPI_ISL_9640051 | BA.1.1 | Positive | Positive | Positive |
| 75 | 31-35 | Male | EPI_ISL_9848001 | BA.1.1 | Positive | Positive | Positive |
| 76 | 46-50 | Male | EPI_ISL_9848002 | BA.1.1 | Positive | Positive | Positive |
| 77 | 76-80 | Female | EPI_ISL_9848003 | BA.1.1 | Positive | Positive | Positive |
| 78 | 81-85 | Male | EPI_ISL_9848004 | BA.2 | Negative | Positive | Positive |
| 79 | 16-20 | Male | EPI_ISL_10067786 | BA.1.1 | Positive | Positive | Positive |
| 80 | 56-60 | Female | EPI_ISL_10067787 | BA.1.1 | Positive | Positive | Positive |
| 81 | 6-10 | Male | EPI_ISL_10067788 | BA.1.1 | Positive | Positive | Positive |
| 82 | 26-30 | Male | EPI_ISL_10067789 | BA.1.1 | Positive | Positive | Positive |
| 83 | 46-50 | Female | EPI_ISL_10067790 | BA.1.1 | Positive | Positive | Positive |
| 84 | 16-20 | Male | EPI_ISL_10067791 | BA.1.1 | Positive | Positive | Positive |
| 85 | 21-25 | Female | EPI_ISL_10067792 | BA.1.1 | Positive | Positive | Positive |
| 86 | 6-10 | Female | EPI_ISL_10067793 | BA.1.1 | Positive | Positive | Positive |
| 87 | 21-25 | Female | EPI_ISL_10067794 | BA.1.1 | Positive | Positive | Positive |
| 88 | 21-25 | Male | EPI_ISL_10067795 | BA.1.1 | Positive | Positive | Positive |
| 89 | 11-15 | Female | EPI_ISL_10067796 | BA.1.1 | Positive | Positive | Positive |
| 90 | 46-50 | Male | EPI_ISL_10067797 | BA.1.1 | Positive | Positive | Positive |
| 91 | 21-25 | Male | EPI_ISL_10115927 | BA.1.1 | Positive | Positive | Positive |
| 92 | 71-75 | Male | EPI_ISL_10115928 | BA.1.1 | Positive | Positive | Positive |
| 93 | 16-20 | Male | EPI_ISL_10334166 | BA.1.1 | Positive | Positive | Positive |
| 94 | 31-35 | Male | EPI_ISL_10334167 | BA.1.1 | Positive | Positive | Positive |
| 95 | 31-35 | Male | EPI_ISL_10334168 | BA.1.1 | Positive | Positive | Positive |
| 96 | 11-15 | Female | EPI_ISL_10334169 | BA.1.1 | Positive | Positive | Positive |
| 97 | 6-10 | Male | EPI_ISL_10334170 | BA.1.1 | Positive | Positive | Positive |
| 98 | 81-85 | Female | EPI_ISL_10334171 | BA.1.1 | Positive | Positive | Positive |
| 99 | 46-50 | Male | EPI_ISL_10334172 | BA.2 | Negative | Positive | Positive |
| 100 | 81-85 | Female | EPI_ISL_10334173 | BA.1.1 | Positive | Positive | Positive |
| 101 | 61-65 | Male | EPI_ISL_10334174 | BA.1.1 | Positive | Positive | Positive |
| 102 | 56-60 | Female | EPI_ISL_10334175 | BA.1.1 | Positive | Positive | Positive |
| 103 | 26-30 | Male | EPI_ISL_10334176 | BA.2 | Negative | Positive | Positive |
| 104 | 71-75 | Male | EPI_ISL_10334177 | BA.1.1 | Positive | Positive | Positive |
| 105 | 41-45 | Female | EPI_ISL_10334178 | BA.1.1 | Positive | Positive | Positive |
| 106 | 11-15 | Male | EPI_ISL_10334179 | BA.1.1 | Positive | Positive | Positive |
| 107 | 36-40 | Female | EPI_ISL_10334180 | BA.1.1 | Positive | Positive | Positive |
| 108 | 6-10 | Male | EPI_ISL_10334181 | BA.2 | Negative | Positive | Positive |
| 109 | 0-5 | Female | EPI_ISL_10730432 | BA.1.1 | Positive | Positive | Positive |
| 110 | 71-75 | Female | EPI_ISL_10730433 | BA.1.1 | Positive | Positive | Positive |
| 111 | 36-40 | Female | EPI_ISL_10730434 | BA.1.1 | Positive | Positive | Positive |
| 112 | 31-35 | Male | EPI_ISL_10730435 | BA.1.1 | Positive | Positive | Positive |
| 113 | 61-65 | Male | EPI_ISL_10730436 | BA.1.1 | Positive | Positive | Positive |
| 114 | 61-65 | Male | EPI_ISL_10730437 | BA.1.1 | Positive | Positive | Positive |
| 115 | 81-85 | Male | EPI_ISL_10730438 | BA.1.1 | Positive | Positive | Positive |
| 116 | 26-30 | Female | EPI_ISL_10730439 | BA.1.1 | Positive | Positive | Positive |
| 117 | 26-30 | Male | EPI_ISL_10730440 | BA.1.1 | Positive | Positive | Positive |
| 118 | 71-75 | Female | EPI_ISL_10730441 | BA.1.1 | Positive | Positive | Positive |
| 119 | 81-85 | Female | EPI_ISL_10730442 | BA.1.1 | Positive | Positive | Positive |
| 120 | 21-25 | Male | EPI_ISL_10730443 | BA.1.1 | Positive | Positive | Positive |
| 121 | 41-45 | Female | EPI_ISL_10730444 | BA.1.1 | Positive | Positive | Positive |
| 122 | 6-10 | Male | EPI_ISL_10730445 | BA.1.1 | Positive | Positive | Positive |
| 123 | 26-30 | Female | EPI_ISL_10730446 | BA.1.1 | Positive | Positive | Positive |
| 124 | 26-30 | Male | EPI_ISL_10730447 | BA.1.1 | Positive | Positive | Positive |
| 125 | 86-90 | Female | EPI_ISL_10981742 | BA.2 | Negative | Positive | Positive |
| 126 | 46-50 | Female | EPI_ISL_10981743 | BA.2 | Negative | Positive | Positive |
| 127 | 46-50 | Female | EPI_ISL_10981744 | BA.2 | Negative | Positive | Positive |
| 128 | 56-60 | Male | EPI_ISL_10981745 | BA.2 | Negative | Positive | Positive |
| 129 | 41-45 | Female | EPI_ISL_10981746 | BA.2 | Negative | Positive | Positive |
| 130 | 6-10 | Male | EPI_ISL_10981747 | BA.2 | Negative | Positive | Positive |
| 131 | 26-30 | Male | EPI_ISL_10981748 | BA.2 | Negative | Positive | Positive |
| 132 | 41-45 | Male | EPI_ISL_10981749 | BA.2 | Negative | Positive | Positive |
| 133 | 56-60 | Male | EPI_ISL_10981750 | BA.2 | Negative | Positive | Positive |
| 134 | 6-10 | Male | EPI_ISL_10981751 | BA.2 | Negative | Positive | Positive |
| 135 | 61-65 | Female | EPI_ISL_10981752 | BA.2 | Negative | Positive | Positive |
| 136 | 51-55 | Male | EPI_ISL_10981753 | BA.2 | Negative | Positive | Positive |
| 137 | 16-20 | Male | EPI_ISL_10981754 | BA.2 | Negative | Positive | Positive |
| 138 | 76-80 | Male | EPI_ISL_10981755 | BA.2 | Negative | Positive | Positive |
| 139 | 76-80 | Male | EPI_ISL_10981756 | BA.2 | Negative | Positive | Positive |
| 140 | 16-20 | Male | EPI_ISL_10981757 | BA.2 | Negative | Positive | Positive |
| 141 | 16-20 | Male | EPI_ISL_11018131 | BA.2 | Negative | Positive | Positive |
| 142 | 0-5 | Male | EPI_ISL_11018132 | BA.2 | Negative | Positive | Positive |
| 143 | 6-10 | Male | EPI_ISL_11018133 | BA.2 | Negative | Positive | Positive |
| 144 | 11-15 | Female | EPI_ISL_11018134 | BA.2 | Negative | Positive | Positive |
| 145 | 51-55 | Female | EPI_ISL_11018135 | BA.2 | Negative | Positive | Positive |
| 146 | 6-10 | Female | EPI_ISL_11018136 | BA.2 | Negative | Positive | Positive |
| 147 | 31-35 | Female | EPI_ISL_11018137 | BA.2 | Negative | Positive | Positive |
| 148 | 36-40 | Male | EPI_ISL_11018138 | BA.2 | Negative | Positive | Positive |
| 149 | 51-55 | Female | EPI_ISL_11018139 | BA.2 | Negative | Positive | Positive |
| 150 | 16-20 | Female | EPI_ISL_11018140 | BA.2 | Negative | Positive | Positive |
| 151 | 61-65 | Male | EPI_ISL_11018141 | BA.2 | Negative | Positive | Positive |
| 152 | 31-35 | Male | EPI_ISL_11018142 | BA.2 | Negative | Positive | Positive |
| 153 | 0-5 | Female | EPI_ISL_11018143 | BA.2 | Negative | Positive | Positive |
| 154 | 11-15 | Male | EPI_ISL_11018146 | BA.2 | Negative | Positive | Positive |
| 155 | 46-50 | Female | EPI_ISL_11050898 | BA.2 | Negative | Positive | Positive |
| 156 | 51-55 | Male | EPI_ISL_11050899 | BA.2 | Negative | Positive | Positive |
| 157 | 41-45 | Female | EPI_ISL_11050900 | BA.2 | Negative | Positive | Positive |
| 158 | 61-65 | Male | EPI_ISL_11050901 | BA.1.1 | Positive | Positive | Positive |
| 159 | 41-45 | Male | EPI_ISL_11050903 | BA.2 | Negative | Positive | Positive |
| 160 | 31-35 | Male | EPI_ISL_11050904 | BA.2 | Negative | Positive | Positive |
| 161 | 41-45 | Female | EPI_ISL_11050905 | BA.2 | Negative | Positive | Positive |
| 162 | 6-10 | Female | EPI_ISL_11050906 | BA.2 | Negative | Positive | Positive |
| 163 | 56-60 | Female | EPI_ISL_11050907 | BA.2 | Negative | Positive | Positive |
| 164 | 6-10 | Female | EPI_ISL_11050908 | BA.1.1 | Positive | Positive | Positive |
| 165 | 0-5 | Male | EPI_ISL_11050909 | BA.1.1 | Positive | Positive | Positive |
| 166 | 36-40 | Female | EPI_ISL_11050910 | BA.1.1 | Positive | Positive | Positive |
| 167 | 51-55 | Male | EPI_ISL_11050911 | BA.1.1 | Positive | Positive | Positive |
| 168 | 41-45 | Male | EPI_ISL_11050912 | BA.1.1 | Positive | Positive | Positive |
| 169 | 31-35 | Female | EPI_ISL_11050913 | BA.1.1 | Positive | Positive | Positive |
| 170 | 11-15 | Male | EPI_ISL_11018144 | BA.2 | Positive | Positive | Inconclusive |
| 171 | 6-10 | Male | EPI_ISL_11018145 | BA.2 | Positive | Positive | Inconclusive |

WGS, whole genome sequencing

Note: SARS-CoV-2 from case #170 and #171 have spike G339N mutation
